# The value of vaccine booster doses to mitigate the global impact of the Omicron SARS-CoV-2 variant

**DOI:** 10.1101/2022.01.17.22269222

**Authors:** Alexandra B Hogan, Sean L Wu, Patrick Doohan, Oliver J Watson, Peter Winskill, Giovanni Charles, Gregory Barnsley, Eleanor M Riley, David S Khoury, Neil M Ferguson, Azra C Ghani

## Abstract

Vaccines have played a central role in mitigating severe disease and death from COVID-19 in the past 12 months. However, efficacy wanes over time and this loss of protection is being compounded by the emergence of the Omicron variant. By fitting an immunological model to population-level vaccine effectiveness data, we estimate that neutralizing antibody titres for Omicron are reduced by 3.9-fold (95% CrI 2.9–5.5) compared to the Delta variant. Under this model, we predict that 90 days after boosting with the Pfizer-BioNTech vaccine, efficacy against severe disease (admission to hospital) declines to 95.9% (95% CrI 95.4%–96.3%) against the Delta variant and 78.8% (95% CrI 75.0%–85.1%) against the Omicron variant. Integrating this immunological model within a model of SARS-CoV-2 transmission, we demonstrate that the size of the Omicron wave will depend on the degree of past exposure to infection across the population, with relatively small Omicron waves in countries that previously experienced a large Delta wave. We show that booster doses can have a major impact in mitigating the epidemic peak, although in many settings it remains possible that healthcare capacity could still be challenged. This is particularly the case in “zero-COVID” countries where there is little prior infection-induced immunity and therefore epidemic peaks will be higher. Where dose supply is limited, targeting boosters to the highest risk groups to ensure continued high protection in the face of waning immunity is of greater benefit than giving these doses as primary vaccination to younger age-groups. In many settings it is likely that health systems will be stretched, and it may therefore be necessary to maintain and/or reintroduce some level of NPIs to mitigate the worst impacts of the Omicron variant as it replaces the Delta variant.

## Introduction

The rapid development and roll-out of vaccines against SARS-CoV-2 has dramatically altered the course of the global pandemic, substantially reducing COVID-19 cases, hospitalisations, and deaths^1–3^. Despite high initial vaccine efficacy against wild-type infection, the emergence of variants of concern, in particular the globally prevalent Delta variant, resulted in a loss of efficacy against mild infection and onward transmission. Recent weeks have seen the emergence of the Omicron variant, exhibiting a large number of mutations in the spike protein gene, with evidence that this variant evades both infection- and vaccine-induced immune responses^4^. Alongside this, there is now clear evidence that vaccine efficacy against existing variants wanes over time^5,6^. Whilst the extent of waning against severe disease (i.e. that requiring hospitalisation) is less than that against mild disease, even small reductions in protection can result in significant rises in hospitalisations and deaths, particularly in high-risk groups. As a result, many countries are now implementing, or considering, booster programmes. Data on the immunogenicity of booster vaccines are encouraging; for third doses administered between 6- and 8-months post dose 2, neutralizing antibody titres (NAT) increased between 4- and 8-fold compared to NATs measured post dose 2^7–9^. Recent data from a clinical trial of the Pfizer-BioNTech vaccine demonstrate a restoration in vaccine efficacy against the Delta variant, reported to be 95% against mild infection^10^. Similar restoration of efficacy is evident in real-world data from Israel and the UK^11,12^. However, the benefit of booster doses in any given population will depend on the current stage of their primary vaccine programme including the supply of vaccine doses, as well as the epidemic that has been experienced to date. Furthermore, the benefits of booster vaccination will depend on the extent to which the Omicron variant replaces the Delta variant and the extent to which the Omicron variant further evades existing immunity. We sought to explore these issues by integrating a model of the dynamics of immunity induced by both infection and vaccines within a population-based virus transmission model.

## Results

To capture the dynamics of vaccine-induced protection, we followed the approach in Khoury et al.^13^ to describe the relationship between NAT, and protection against mild and severe disease, over time. More recent data on the decay of NAT over time after a two-dose vaccination regimen suggest an initial rapid decline (up to 90 days^14^, in line with the 108-day estimate in^13^ with a second slower phase of decay with a half-life of approximately 500 days^15^. We further re-parameterised the model to incorporate data on NAT and vaccine efficacy following dose 1 from clinical trials^16–18^, and on the immunogenicity of booster doses delivered between 6- and 8-months post dose 2^19^. To capture reduced efficacy against the Omicron variant, we applied a multiplicative scaling factor to reflect reduced neutralization of this variant for all vaccines modelled. Reductions in NATs for previous variants compared to wild type (WT) have been in the range 1.6–8.8-fold^20^ whilst early studies have shown between a 6- to 40-fold reduction in NAT relative to WT for the Omicron variant^21–23^. We refer to this reduction as the variant fold reduction (VFR).

We used these data to generate priors (see Table S1) to fit the model to real-world estimates of vaccine efficacy against mild disease (PCR+ tests which include symptomatic cases and asymptomatic infections detected through screening in schools and workplaces), severe disease (hospitalisations) and death (within 28 days of a positive test) with the Delta and Omicron variants from England^24,25^. Data were available for two vaccine regimens: Pfizer-BioNTech primary series with Pfizer-BioNTech booster (PF-PF) and Oxford-AstraZeneca primary series with PF booster (AZ-PF). As demonstrated in the Phase III trials and subsequent population studies for the Delta variant, both vaccine regimens are estimated to generate initial substantial protection against mild and severe disease (Figure 1). The model also captures the observed slower waning of efficacy against severe disease compared to the faster waning against mild disease, generated by the different dose-response curves fitted to these two endpoints (Figure 1, Figure S1). For these vaccine regimens, we estimate that the boosting with PF (used in both regimens) increases NATs by 1.5-fold (95% credible interval, CrI 1.4–1.8) compared to NATs following dose 2 of PF and 3.0-fold (95% CrI 2.5– 3.9) compared to NATs following dose 2 of AZ, resulting in similar NATs following the boost regardless of the primary course. We estimate a 3.9-fold (95% CrI 2.9–5.5) reduction in NATs against the Omicron variant compared to the Delta variant (20) which translates to approximately a 10–23-fold drop in NAT compared to WT, consistent with a meta-analysis of NAT studies^26^. This drop in NATs matches the observed drop in vaccine efficacy against mild disease. Under this model, we would expect to see a similar drop in vaccine efficacy against severe disease and death, although additional, more broadly based immunological mechanisms may provide protection above those indicated by NAT levels. Furthermore, we expect to see a tail in this distribution meaning a minority of people who exhibit a poor response to vaccination could be at particularly high risk of severe outcomes (Figure 1, illustrated as individual trajectories).

**Figure 1:**
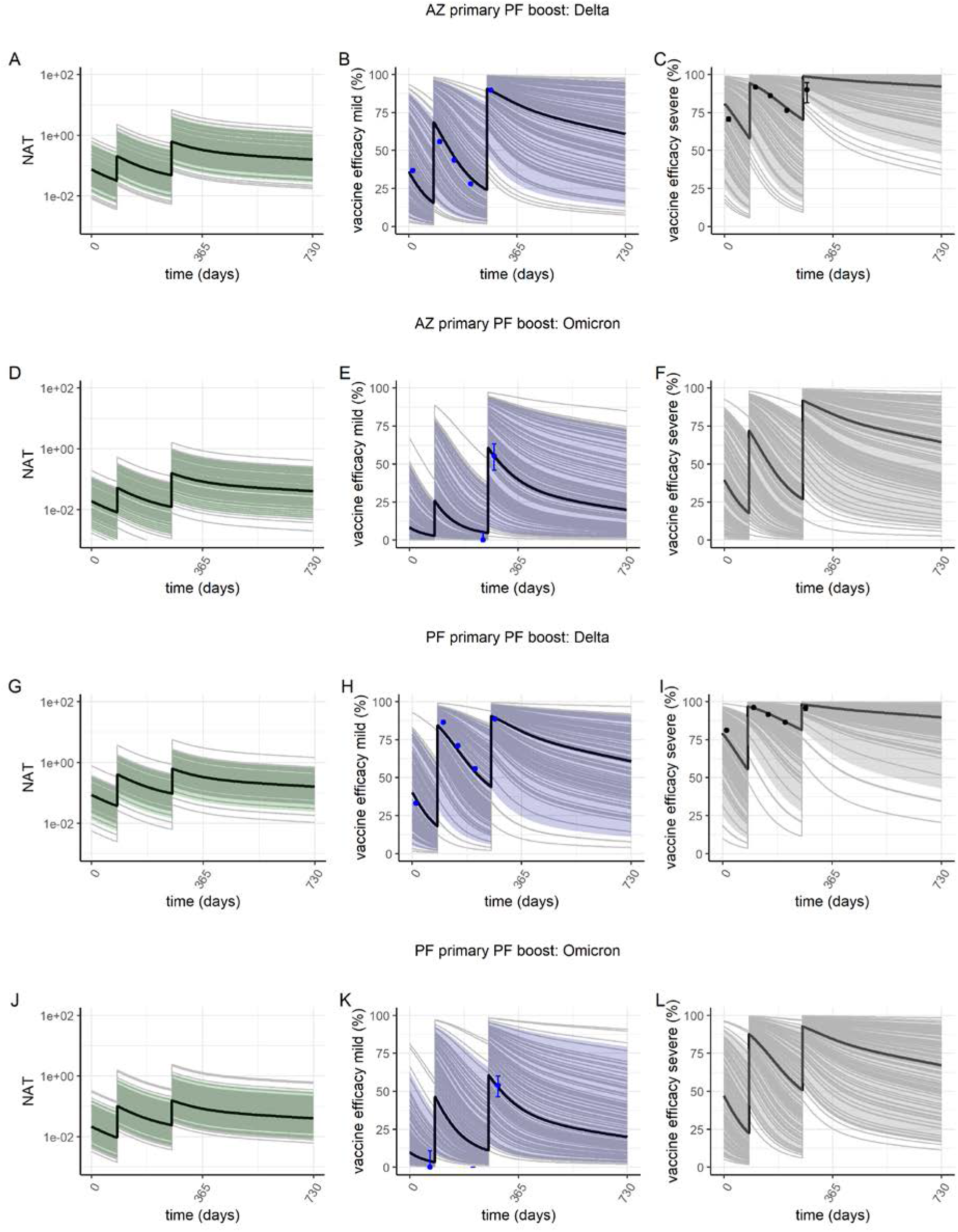
Predicted vaccine efficacy over time. Plots show neutralizing antibody titre (NAT) (green), vaccine efficacy against mild disease (purple) and severe disease (grey) for the AZ-PF regimen (panels A-F) and PF-PF regimen (panels F–L). The solid black line shows the posterior median estimate, colour bands the 95% range of individual responses and grey lines show 100 individual draws from the variation estimated between individuals in *(13)* illustrating the impact of variability in immune responses across the population. In panels B, E, H and K, the blue points are estimates of vaccine efficacy against mild disease (i.e. PCR+ tested infections which include asymptomatic infections detected through regular screening in schools and some workplaces) with the Delta and Omicron variants in England from *(24, 25)*. In panels C and I, the black points represent vaccine efficacy against hospitalisation with the Delta variant from the same source. Panels A–C and G–I show NAT and vaccine efficacy against the Delta variant; panels D–F and J–L show NAT and vaccine efficacy against the Omicron variant.

Table 1 shows the predicted vaccine efficacy against mild disease, severe disease and death following the primary series and a booster dose for these two regimens. Our results suggest that vaccine efficacy against both mild and severe disease could be substantially impacted by the Omicron variant. Under this model, we predict that 90 days after PF boosting, efficacy against severe disease will decline to 95.9% (95% CrI 95.4%–96.3%) against the Delta variant and 78.8% (95% CrI 75.0%–85.1%) against the Omicron variant. In making this projection we assume that there is a shift in the decay rate following boosting with the period of faster decay halved to represent a higher proportion of long-lived antibody-secreting cells (ASCs) compared short-lived ASCs that would result in continuous antibody production over a longer period (sensitivity to this assumption is given in Table S3). Whilst there remains considerable uncertainty as to the level of protection and its longevity post-boost, our scenarios suggest that either repeated boosts and/or Omicron-variant specific vaccines will likely be needed to restore protection against severe disease to the levels seen in Phase III vaccine trials^25^.

**Table 1:**
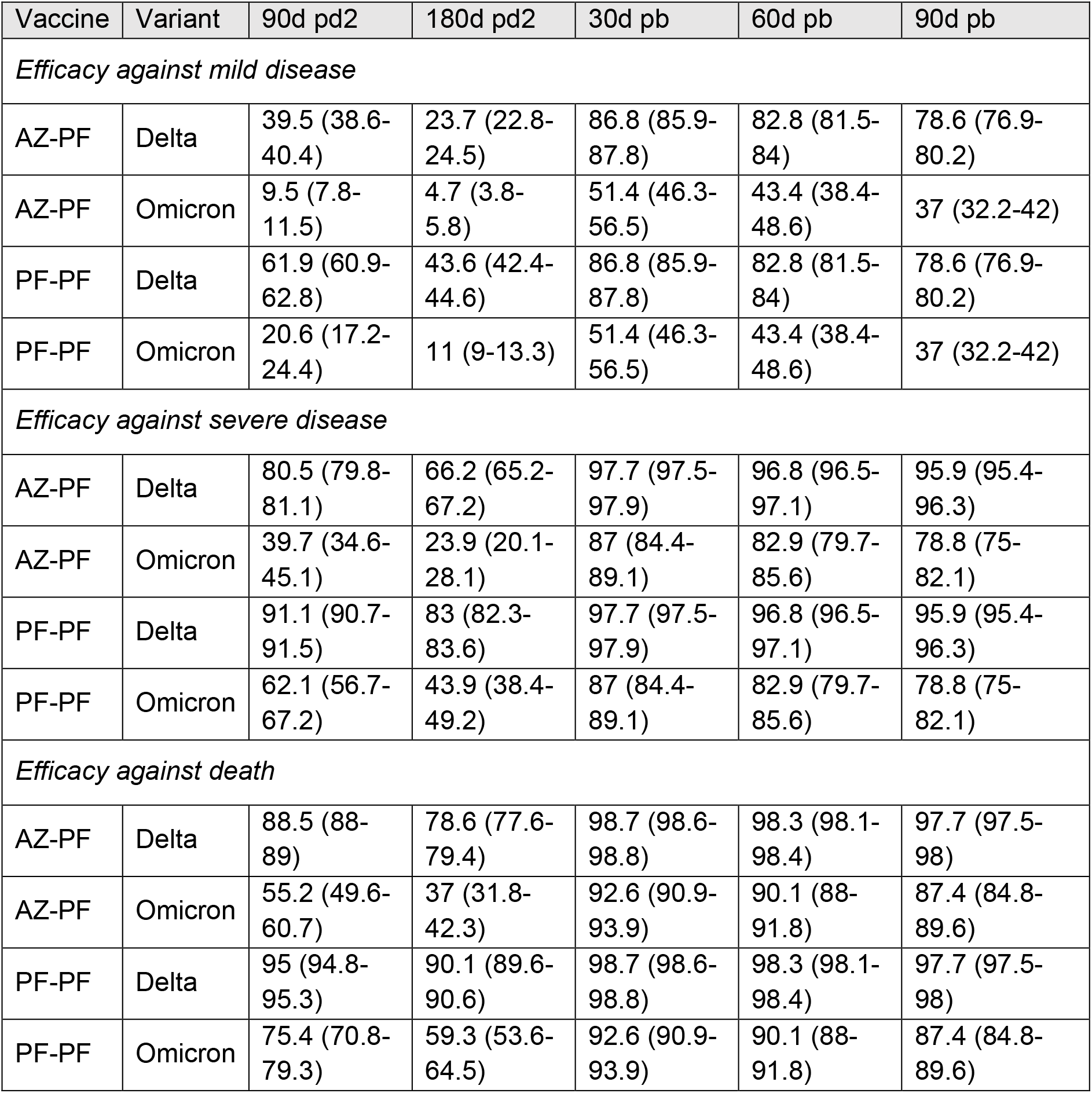
Estimated vaccine efficacy against mild disease, severe disease and death for the AZ-PF and PF-PF vaccine regimens as a function of time since dose 2 or booster. Estimates are shown for the Delta variant and the Omicron variant. Values shown are the posterior median and 95% credible intervals. pd2 = post dose 2; pb = post booster.

We used the same approach to capture infection-induced immunity and its interaction with vaccine-induced immunity. Cohort studies in high-risk populations suggest that protection against mild infection is consistently maintained at levels above 90% for the first 8–12 months post infection^27–31^, likely due to the broader immunological response induced by the whole virus compared to the relatively limited response induced by the PF and AZ vaccines that elicit responses against only the spike protein. We capture this by calculating the level of NAT acquired after an infection that would generate 90% mean protection over 1 year under our model. We assume that infection-induced NAT decays at the same rate as that induced through booster dose vaccination (i.e. at a slower rate than following primary vaccination with the period of faster decay halved compared to the rate of decline following primary vaccination). We find that a NAT of 1.7 relative to the level measured in convalescents is required to match 90% protection from re-infection over 1 year (consistent with a recent modelling study^32^). The net result is therefore that we predict a slower decline in protection in those previously infected compared to the protection afforded through vaccination. We assume that infection-induced NAT to the Omicron variant is reduced by the same VFR as vaccine-induced NAT. For our estimated fold-reduction of vaccine-induced immunity to the Omicron variant compared to the Delta variant, this would imply 60% (95% CrI 50–68%) mean protection against infection from the Omicron variant over one year and 90% mean protection against hospitalisation (95% CrI 85-93%) in those previously infected with another variant, consistent with levels reported from South Africa and the UK^25,33^.

In many countries, vaccination is occurring in populations that have already been exposed to past waves of infection and will have substantial levels of infection-induced immunity. Studies of the immunogenicity of the PF vaccine in previously infected individuals suggest that a single vaccine dose generates a response similar to or exceeding two vaccine doses in infection naïve individuals^34,35^. This effect is captured in our model, with the first vaccine dose increasing NAT from prior infection to levels equivalent to those seen post dose 2 in uninfected individuals.

For both infection- and vaccine-induced immunity, longer-term waning of NAT depends on the rate of loss of long-lived antibody secreting cells. This is captured in our model by the second slower decline, although this rate of decline is uncertain. It is plausible that the degree of waning after infection is slower than that after vaccination, and therefore that high prior levels of infection-induced immunity could provide a higher degree of protection than captured here. It is also likely that aspects of the overall immune response will vary by age; however, there are currently insufficient data to explore the impact of age on waning vaccine efficacy or immune escape from booster doses due to the shorter follow-up in younger populations.

We next considered how booster doses should best be deployed given the current state of the global pandemic. To do so, we developed an individual-based model of SARS-CoV-2 transmission within which we embedded the immunological model (Figure S2, Figure S3). The model is parameterised to capture differences between countries in demography, age-mixing patterns, and health system impacts on access to hospital facilities^36,37^. We additionally sought to capture characteristics of the Omicron variant based on emerging data. Exponential growth of the omicron variant is being experienced in many countries. To estimate the intrinsic transmissibility of the Omicron variant associated with our estimated level of immune escape, we compared model-generated scenarios for the UK and Australia to the early growth rate of the epidemics, choosing these two populations as illustrative of differing prior exposure to the Delta variant. For the UK we find that the growth rate of ∼0.07 per day is consistent with model trajectories with Omicron Rt∼3.5 (Figure S4A). For Australia we find that the more rapid growth rate of ∼0.14 per day is consistent with a slightly lower Omicron Rt∼3 (Figure S4B). The levels of Rt experienced depend on population behaviour alongside rates of vaccination and the degree of prior infection. However, given the level of immune escape that we estimate from the vaccine efficacy data, the intrinsic transmissibility appears similar to estimates for Delta (although the same pattern could be explained by changes in the generation time for Omicron compared to Delta). For our central scenarios we therefore kept the Omicron Rt at the same level as the Delta Rt, and additionally explored “best-case” and “worst-case” scenarios with reduced and increased transmissibility respectively.

We further incorporated recent evidence from the UK suggesting intrinsic lesser severity of infections with the Omicron variant^38–42^, reducing the probability of hospitalisation by 50% (range 40%–70%) and the probability of subsequent admission to intensive care by a further 58% (conditional on hospitalisation), which results in a 71% (range 65%–83%) reduction in admission to intensive care compared to the Delta variant.

To capture the different epidemic trajectories that have occurred alongside access to vaccines, we stratify the current epidemiological state of countries into three broad categories. The first exemplar setting captures countries that have experienced substantial past transmission (and hence have a substantial level of infection-induced immunity) and that also have a high level of access to vaccines. Many high- and upper-middle-income countries fall into this category – including North America, countries in Central/South America, the Middle East and much of Europe. We created a representative epidemic profile for such countries, with a first wave occurring between March and May 2020, a second wave occurring during the northern hemisphere winter of 2020/21, and transmission gradually increasing (i.e. interventions being relaxed) from mid-2021 (Figure S5). We assumed that the primary courses of vaccines (using PF-PF vaccine schedule properties) were administered from January 2021 onwards at a constant rate of 5% of the population dosed per week, a pace that would result in approximately 80% of the population being vaccinated by the end of August 2021 (Figure 2A). Whilst patterns of roll-out have varied between countries, most prioritised the elderly and high-risk individuals initially. We therefore mimicked this prioritisation approach, with the oldest individuals vaccinated first and vaccines delivered sequentially until everyone aged 10 years or above is vaccinated. Throughout we assumed 90% uptake within each age-group.

**Figure 2:**
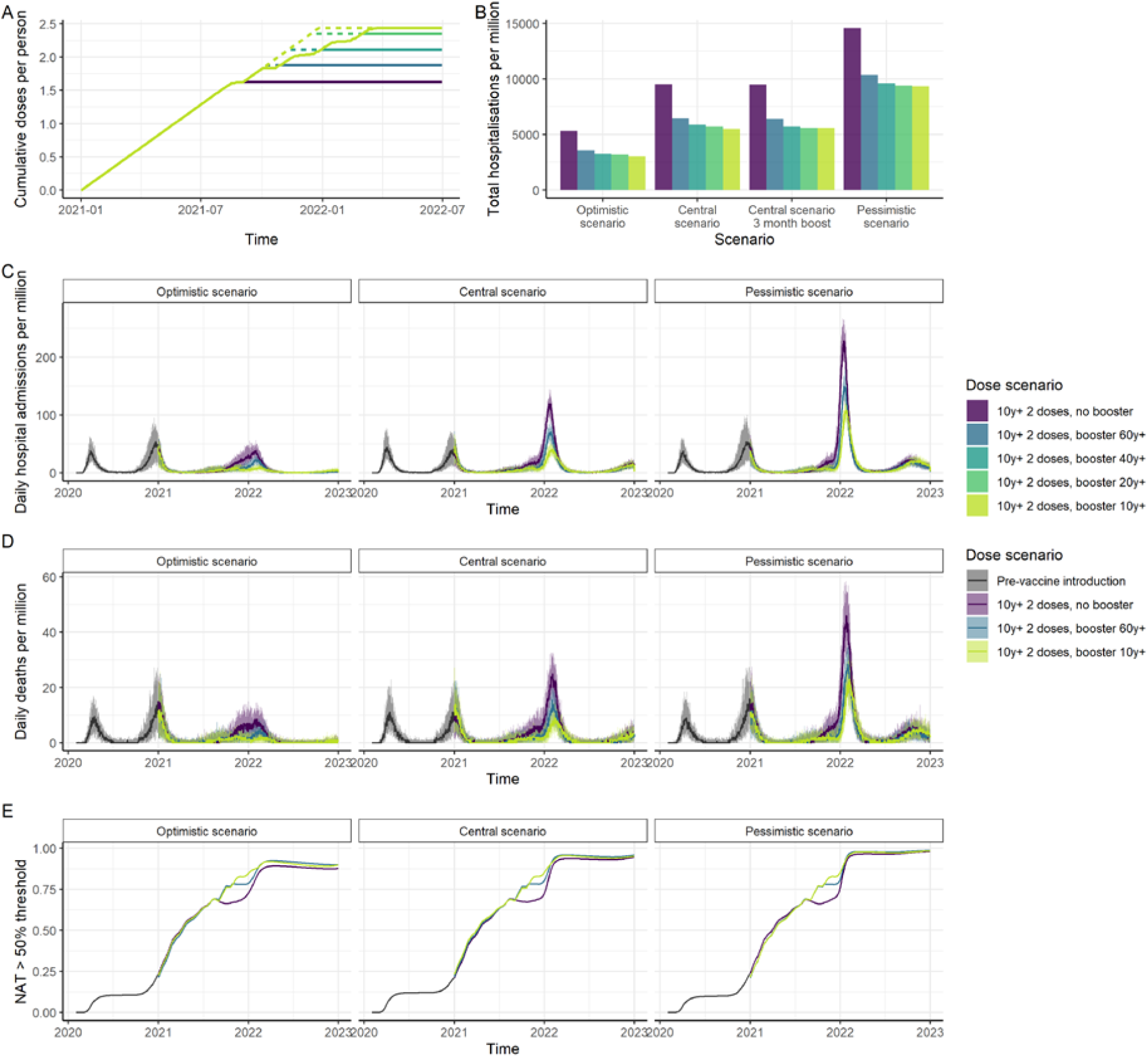
Impact of vaccination in a high-income country setting with substantial prior transmission and high vaccine access. (A) Cumulative vaccine doses delivered per person over time. Solid lines represent a 6-month period between dose 2 and the booster dose; dashed lines represent a 3-month period. Vaccine impact is shown for five dose scenarios: no booster doses, booster doses to those aged 60+, booster doses to those aged 40+, booster doses to those aged 20+ and booster doses to those aged 10+. Impact is shown for three epidemiological scenarios: central, with our central estimate of the VFR (3.9), Omicron Rt=3.5 (the same as Delta) and the risk of hospitalisation reduced by 50% for Omicron compared to Delta; optimistic with our lower estimate of VFR (2.9), Omicron Rt=3.25 and risk of hospitalisation reduced by 70%; and pessimistic with our upper estimate of the VFR (5.5), Rt=3.75 and risk of hospitalisation reduced by 40%. All scenarios assume 90% vaccine uptake in each group and the risk of death further reduced by 58% from the risk of hospitalisation. (B) Total hospitalisations per million post vaccine introduction at the beginning of 2021 through to end-2022, for the central, best-case and worst-case scenarios, and assuming a 6-month period between dose 2 and the booster, except where specified. (C) Daily hospital admissions; and (D) Daily deaths per million population for the best-case, central, and worst-case scenarios. (E) Proportion of the population with NAT higher than the titre relative to convalescent required to provide 50% protection from infection, for the central VFR estimate.

In countries in which the adult population has already received their primary (2 dose) immunisation, we find that administration of a booster dose (6 months post dose 2) to those aged 60+ years has a substantial impact, reducing hospitalisations and deaths by 32 and 34% respectively from January 2021 to end-2022 compared to not boosting and assuming no other NPIs are in place (Figure 2B–D). Due to the timing of the predicted Omicron wave relative to vaccination (Figure 2A), the impact of progressively continuing the booster programme into younger age groups will depend on the speed of vaccination coupled with how transmission evolves in 2022, although infections would likely be reduced (Figure S6).

The size of the Omicron peak in terms of admissions to hospital and deaths compared to the previous waves depends on the balance between the increased transmissibility and decreased severity. Across our range of scenarios for the impact of the emergence of the Omicron variant and in the absence of a booster programme we predict a significant winter wave at levels comparable to or exceeding previous waves (Figure 2C–D) with hospitalisations peaking at between 45 - 220 per million per day and deaths at 15 - 45 per million per day, across a range of values for the VFR, transmissibility and hospitalisation risk of the Omicron variant relative to the Delta variant (Figure 2). To mitigate this, high coverage of booster doses is required, particularly in the highest-risk age-groups. With booster doses for the 60+ age-group, and assuming 90% uptake, we expect a peak in hospitalisations that in our optimistic and central scenarios is lower than previous waves whilst higher in our most pessimistic scenario, with hospitalisations peaking at between 40–150 per million per day and deaths at 10–30 per million per day across all three scenarios (Figure 2C–D). With booster doses rolled out across the population (10 years and above), hospitalisations peak at 40-110 per million per day and deaths at 10-25 per million per day. This reduction in the peak and overall impact compared to previous waves is in part due to relatively high levels of pre-existing immunity from prior infection (Figure 2E, Figure S7) and hence these levels will vary across settings depending on their past epidemic. Against this, the booster programme is predicted to ensure that a large proportion of the population have a high level of protection by the beginning of 2022 (Figure 2E, Figure S7). Nevertheless, it may still be necessary to maintain and/or reintroduce some NPIs to flatten the peak of the Omicron variant as it replaces the Delta variant in order to reduce the pressure on health services. Notably, given the high immune escape of the Omicron wave and hence widespread exposure of the population to infection, we expect very high levels of immunity across the population to be retained through 2022 (Figure 2E, Figure S7).

The second set of countries are those that have experienced substantial prior transmission (and hence have a substantial level of infection-induced immunity) but that to date have had limited access to vaccines. Many low- and lower-middle-income countries fall into this category (though we note that several LMIC have successfully limited transmission and so are more comparable to our third category of countries). In these settings, decisions on future vaccine roll-out are likely to be constrained by limited vaccine supply and/or uptake. To assess the value of boosters we therefore compared two roll-out scenarios. In the first, the primary immunisation (assuming two AZ doses) is delivered to all individuals in the highest risk groups (40+ years) and is then paused before delivering AZ boosters to these same groups at 6 months following dose 2 (for which we assume the same scaling for a booster dose relative to dose 2 as estimated for PF). In the second scenario, we assume that the same number of vaccine doses is instead used to continue primary vaccination into younger age-groups (Figure 3A). The latter results in those aged 30+ years being targeted for two vaccine doses. These vaccine roll-out scenarios were evaluated against a similar background epidemic explored in high-income settings (Figure S5), but with fewer NPIs in place during 2021, a slower pace of vaccine roll-out and lower coverage as a result of limited supply. We find that prioritising limited vaccine supply for boosters in the elderly higher-risk population has a greater public health impact, reducing hospitalisations and deaths by 8% and 9% respectively compared to using these same doses to immunise younger age-groups in an effort to reduce transmission (Figure 3B, C, D). Total infections were similar between the two targeting approaches (Figure S8). Similar results are obtained if the booster cut-off was chosen as 60+ years rather than 40+ years, with lower overall impact due to the smaller number of doses delivered under this scenario (reducing hospitalisations by 3% and deaths by 5% if switching to boosters compared to vaccinating younger age-groups, Figure S9). The exception is where a slower rollout results in booster vaccination occurring after the lifting of NPIs, “missing” the epidemic wave (Figure 3D).

**Figure 3:**
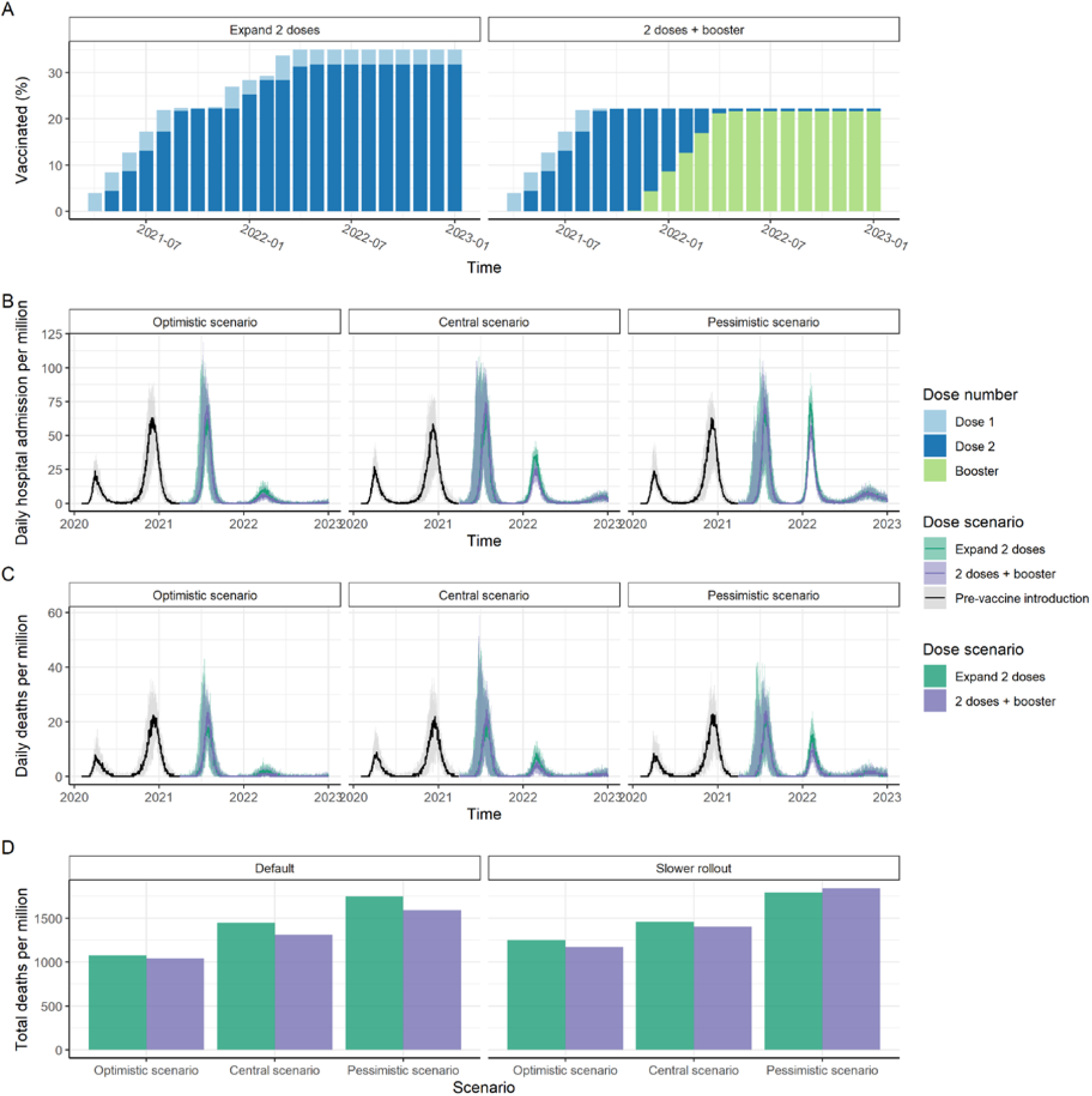
Impact of vaccination in a low-middle-income country setting with substantial prior transmission and low vaccine access, where individuals 40+ years are initially targeted. Two strategies for distributing a limited vaccine supply are shown (assuming AZ is used for both the primary and booster doses). In the first (“Expand 2 doses”), no booster doses are administered, and the supply is therefore delivered to a wider proportion of the population. In the second (“2 doses + booster”) the same supply is delivered to the 40+ age-group (2 dose primary immunisation and booster dose 6 months post dose 2) and no younger groups receive the primary series. (A) Cumulative proportion of the population receiving dose 1 (light blue), dose 2 (dark blue) and the booster (green) each month. (B) Daily hospital admissions; and C) Daily deaths per million population for our central, best-case and worst-case scenarios (see legend to Figure 2). Total deaths per million population from the start of vaccination in 2021 to end-2022 for (D) the three scenarios with the default and slower roll-out rate. Impact is shown for three epidemiological scenarios: central, with our central estimate of the VFR (3.9), Omicron Rt=4 (the same as Delta in this setting) and the risk of hospitalisation reduced by 50% for Omicron compared to Delta; optimistic with our lower estimate of VFR (2.9), Omicron Rt=3.75 and risk of hospitalisation reduced by 70%; and pessimistic with our upper estimate of the VFR (5.5), Rt=4.25 and risk of hospitalisation reduced by 40%. All scenarios assume 80% vaccine uptake in each group and the risk of death further reduced by 58% from the risk of hospitalisation. Total deaths are shown for the default vaccine rollout (2% of the population per dose per week) and booster dose scenario (180 days post dose 2, “Default”), as well as assuming a slower rollout (1% per week, “Slower rollout”). Results for the scenario where the 60+ years population is initially targeted are shown in **Error! Reference source not found**.

Despite the slower pace of vaccine roll-out that has occurred to date in these countries, the Omicron variant is not predicted to generate as large an epidemic as in previous waves (Figure 3B, C) due to widespread infection-induced immunity consistent with the high levels of seroprevalence reported following the Delta wave (Figure 3D). Under our central scenario, and assuming roll-out of primary immunisation to younger age-groups, we expect a reduction in peak hospitalisations of 49% and in deaths of 59% compared to the preceding Delta wave. Under our best-case scenario, hospitalisations will be reduced by 82% and deaths by 86%. These scenarios are in line with reports from South Africa^41^. Only under the worst-case scenario would we expect to see peaks in hospitalisation of similar magnitude to the Delta wave, and even here deaths are predicted to be 29% lower. As in other settings, the precise level of the peak is sensitive to assumptions regarding the severity of the predominant circulating variant (Figure 3E) as well as levels of past infection.

The final set of countries are those that successfully interrupted transmission (the so-called “zero-COVID” countries, mostly in east Asia and the Pacific) and therefore have limited infection-induced immunity. These countries face a different challenge, aiming to gradually reopen their borders and their economies once vaccine coverage is high enough to limit the health impact of increased transmission. In these countries, if NPIs had been relaxed completely once 80% of the adult population had received their primary immunisation (assuming two PF doses, in these simulations representing September 2021), it is likely that they would have experienced a significant Delta wave despite high vaccine coverage that would subsequently be followed by an Omicron wave (Figure 4, Figure S11). Relaxing NPIs following delivery of booster doses to the highest risk 60+ age-group (in these simulations representing November 2021) is still likely to generate a large epidemic (comparable to the waves experienced in other countries), with the introduction of the Omicron variant generating a tail in the epidemic (Figure 4). The impact of delaying NPI relaxation until the population is fully boosted (here illustrated with an April 2022 relaxation) is more uncertain. By this time vaccine efficacy following the booster dose will have waned in the highest-risk groups, and hence under our central scenario we observe a larger epidemic wave (Figure 4). However, under our optimistic scenario with less immune escape, lower transmissibility and lower severity, we observe a much smaller epidemic wave (Figure S12). If NPIs are relaxed more gradually alongside the scale-up of booster doses across the population, the peak of this epidemic, and hence demand on health services, can be mitigated although the cumulative impact depends on the characteristics of the Omicron variant and the durability of the booster dose. As countries such as Australia are now experiencing, given the lack of infection-induced immunity in the population coupled with reduced vaccine efficacy against the Omicron variant, levels of demand on healthcare could stretch healthcare capacity, with peak daily hospital admissions at levels that far exceed the levels these countries have previously experienced in the pandemic. These countries will therefore need to carefully consider how well protected their vaccinated at-risk populations are when opening up given the lack of infection-induced immunity across the population.

**Figure 4:**
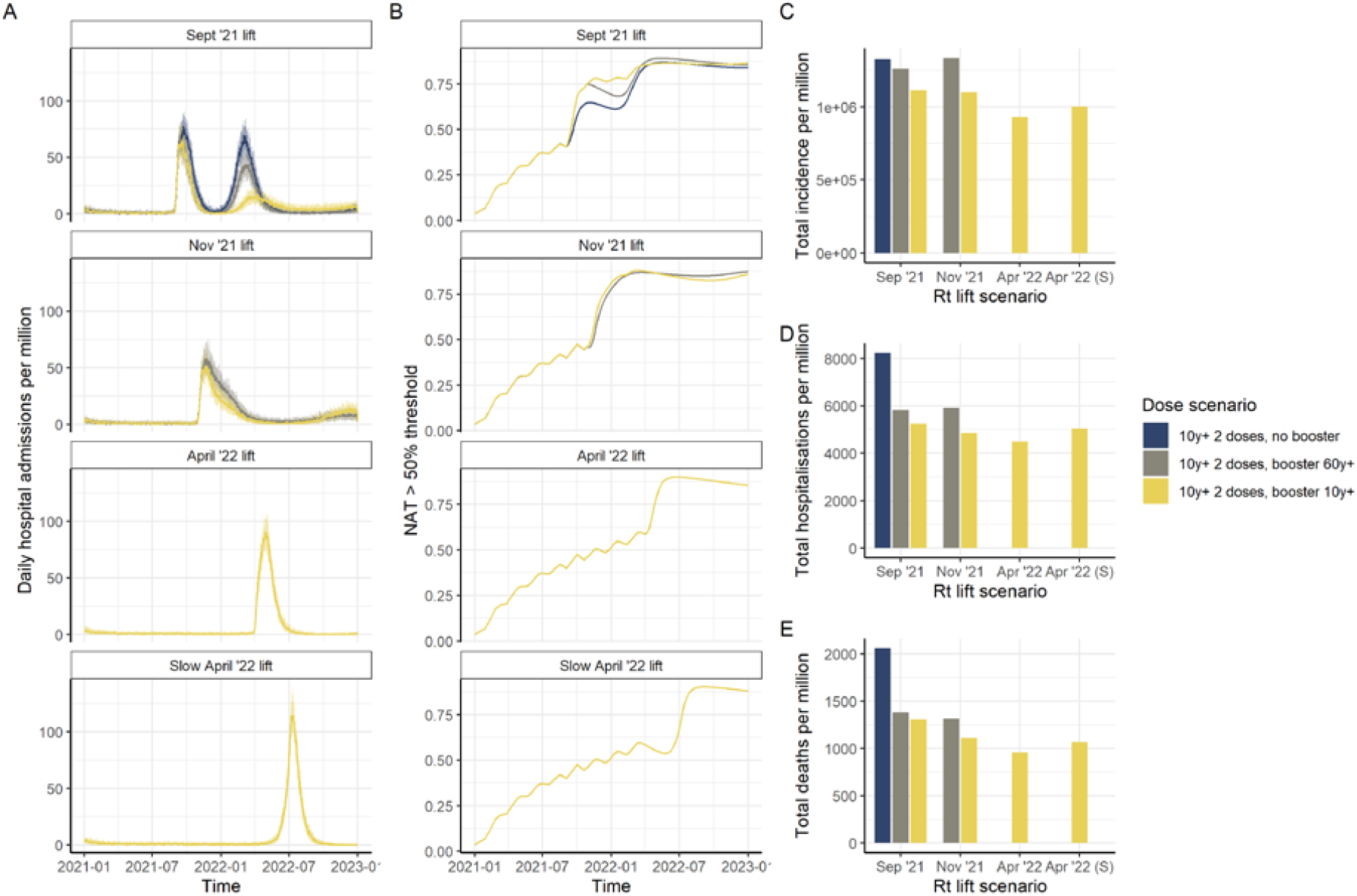
Impact of vaccination in a high-income country setting with minimal prior SARS-CoV-2 transmission and high vaccine access. (A) Daily hospital admissions per million population; and (B) Proportion of the population with NAT higher than the titre relative to convalescent required to provide 50% protection from infection, for the scenario where 80% of the population is immunised (2 doses) before NPIs are lifted (i.e. R_t_ increases). Following Rt increasing (“Sept ‘21 lift”), vaccination continues at a population dose rate of 5% per week until 90% uptake is achieved in the 10+ years population (brown); boosters are administered to the 60+ years population 6 months post dose 2 (dark blue); or boosters are administered to the 10+ years population (yellow). Additional panels show the impact of lifting NPIs after boosters are given to 60+ years (“Nov ‘21 lift”) or after boosters are given to 10+ years (“April ‘22 lift” representing a lifting over a one-month period; “Slower April ‘22 lift” representing lifting over a four-month period). The central VFR estimate, a maximum transmission of R_t_ = 3 (based on the early growth rate of Omicron in Australia) are applied. We assume a 50% reduction in the risk of hospitalisation with Omicron compared to Delta and a further 58% reduction in the risk of death. Panels C, D, and E show the total incidence, hospitalisations, and deaths respectively, per million population from the start of vaccination in 2021 to end-2022, for each scenario.

## Conclusions

With the rapid spread of the Omicron variant the Delta variant has already been replaced in many countries and this trend is likely to continue globally. Emerging immunogenicity data clearly point to substantial reductions in NAT whilst preliminary vaccine efficacy estimates demonstrate a substantial reduction in protection from infection. Our estimates suggest that this is likely to translate into small but important reductions in efficacy against severe disease and death, although this may be moderated by lower VFR for Omicron compared to other variants and increased longevity of T cell-mediated immunity compared to NAT. A further remaining uncertainty is how severe the disease caused by the Omicron variant is compared to disease caused by previous variants. Estimates to date from South Africa, Canada and the UK^38–42^ show a considerable reduction. However, across these ranges whilst the early signs are that the impact is likely to be less severe than the Delta variant, the impact will depend on the population age-distribution, the level of vaccination coverage and the extent of prior waves of infection. Whilst it may take several weeks to fully understand the severity of Omicron infections, governments need to consider putting mitigations in place now to alleviate any potential impact. Our analyses demonstrate the importance of delivering booster doses as part of the wider public health response to restore vaccine efficacy against the Omicron variant. Prioritising these boosters to high-risk and older populations over primary vaccination in younger age-groups should be part of this response if vaccine dose supply is limited or vaccine roll out is delayed.

## Supporting information

Supplementary Methods and Results

## Data Availability

All data produced in the present study are available online on the Github link provided in the manuscript.

https://mrc-ide.github.io/safir

https://github.com/mrc-ude/global_covid_vaccine_booster_paper.

## Acknowledgements

We thank Bob Verity, Nick Grassly, Sarah Pallas and the WHO SAGE working group on COVID vaccines for helpful comments and suggestions on earlier parts of this work.

## Author contributions

Conceptualization and study design: ACG, ABH, SLW, OJW, GB, PW; Individual-based model development: SLW, GC, OJW; Vaccine efficacy model fitting: NMF, PD, ABH, DK, ACG; Analysis: ABH, ACG, PD, SLW; Visualization: ABH, ACG, PD, SLW; Writing – original draft: ACG, ABH, SLW, EMR; Writing – review & editing: All authors

## Data and materials availability

The model code is open access at https://mrc-ide.github.io/safir. All analysis code is available at https://github.com/mrc-ude/global_covid_vaccine_booster_paper.

