## Supplementary Methods and Results for "The value of vaccine booster doses to mitigate the global impact of the Omicron SARS-CoV-2 variant"

Supplementary Materials

### Methods

#### *Immunological Model*

To capture the dynamics of vaccine-induced and infection-induced protection, we followed the approach of Khoury et al. (*13*) by considering the relationship between neutralizing antibody titres (NAT) over time, and protection against mild disease (which we assume to be the same as protection against any infection) and against severe disease requiring hospitalisation. We first model each individual's NAT over time, which is based on a biphasic exponential decay where $n_{ij}$ is the initial NAT of vaccine *i* drawn from a $\log_{10}$-normal distribution at dose *j*. We assume an initial period of fast antibody decay with decay rate $\lambda_1 = -ln(2)/hl_s$, representing the combined biochemical decay of antibodies and the ongoing production of antibodies by circulating plasma cells, followed by a second period of slow decay with decay rate $\lambda_2 = -ln(2)/hl_l$, representing the long-lived plasma cells. This is represented by the bi-phasic exponential function:

$$n(t) = n_{ij}\frac{exp(\lambda_1 t + \lambda_2 t_s) + exp(\lambda_2 t + \lambda_1 t_s)}{exp(\lambda_1 t_s) + exp(\lambda_2 t_s)}$$

where $t_s$ is the period of switching between the fast and slow declines.

We assume a logistic relationship between NAT and efficacy to capture time-varying vaccine protection against infection, severe disease and death ($E(t)$), given by the function

$$E(t) = \frac{1}{1 + e^{-k[log_{10}(n(t)) - log_{10}(n_{50})]}},$$

where $k$ is the fitted shape parameter and $n_{50}$ is the NAT relative to convalescent NAT required to provide 50% protection (from infection, severe disease, or death).

Khoury *et al* (*13*) obtained parameters for this model of vaccine-induced protection by fitting to efficacy data from Phase III trials, using NAT data following dose 2 which are based on the mean 28-day values reported in clinical trials relative to the mean titre for a convalescent individual. By definition, on this scale, the NAT induced by infection in convalescent individuals is 1.

However, the parameters from Khoury *et al*. (*13*) are based on trials conducted during the early period of the pandemic and prior to the emergence of variants. To obtain appropriate estimates for the Delta and Omicron variants and to better inform the parameters determining waning, we re-fitted the model to data on vaccine efficacy from a population-wide study in England (*24*, *25*). This study provides estimates of vaccine efficacy for two vaccine regimens over time – Pfizer-BioNTech primary and Pfizer-BioNTech boost, and Oxford-AstraZeneca primary with Pfizer-BioNTech boost – based on data linkage generating a whole population cohort.

We fitted the model to the estimated vaccine efficacies against the two vaccine regimens and for the Delta and Omicron variants using Bayesian methods and assuming a Binomial likelihood for the vaccine efficacy estimates. We used prior estimates from the original model papers to inform our fitting.

To obtain prior estimates for the impact of a booster dose, we collated reported data on immunogenicity from booster trials undertaken between 3- and 8-months following dose 2

(*19*, *43*). Across these studies, the level of the boost relative to dose 2 NAT differed depending on the vaccine and the VOC. For wild-type, the studies reported a 4–5-fold boost after the booster dose compared to dose 2. However, for the Beta variant this appears to be 10–20 fold higher, whilst for Delta the boost was in the range 5–7 fold compared to dose 2. We thus used a 6-fold boost a prior mean compared to dose 2.

Our priors and their sources are summarised in **Table S1**. Dose parameters and titre parameters were transformed to the log10 scale, all other parameters were fitted on a linear scale. We used Normal distribution priors on these scales with mean and standard deviations shown in **Table S1**. Model fitting was undertaken using MCMC methods in the Dr.Jacoby R package (https://mrc-ide.github.io/drjacoby/index.html).

**Table S1: Prior and posterior parameter estimates for the immunological model**

| Parameter | Symbol | Prior Mean | Prior S.D. | Posterior Estimate – median (95% credible interval) | Reference for Prior Mean |
|---|---|---|---|---|---|
| NAT against Delta for dose 1 of the AstraZeneca vaccine relative to convalescent | $n_{AZ,1}$ | 0.5* $n_{AZ,2}$ | 0.5 | 0.075 (0.040, 0.135) | - |
| NAT against Delta for dose 2 of the AstraZeneca vaccine relative to convalescent | $n_{AZ,2}$ | 32/59 for WT from data in (*13*) With fold-reduction applied | 0.25 | 0.205 (0.118, 0.352) | (*13*, *20*) |
| Fold-reduction for Delta relative to WT for AstraZeneca vaccine | - | 3.9 | 1.5 | 2.6 (1.5, 4.6) | (*20*) |
| NAT for dose 1 of the Pfizer vaccine against Delta | $n_{PF,1}$ | 0.5* $n_{PF,2}$ | 0.5 | 0.086 (0.047, 0.154) | - |
| NAT for dose 2 of the Pfizer vaccine against Delta | $n_{PF,2}$ | 223/94 from data in (*13*) With fold-reduction applied | 0.25 | 0.404 (0.235, 0.704) | (*13*, *20*) |
| Fold-increase in NAT for booster dose of Pfizer vaccine against Delta compared to dose 2 of Pfizer | $n_{PF,3}$ | 6 | 4 | 1.5 (1.4, 1.8) | (*19*, *43*) |

| | | | | | |
|---|---|---|---|---|---|
| Fold-increase in NAT for booster dose of Pfizer vaccine against Delta compared to dose 2 of AstraZeneca | $n_{AZ,3}$ | 6 | 4 | 3.0 (2.5, 3.9) | (*19*, *43*) |
| Fold-reduction for Delta relative to WT for Pfizer vaccine | - | 3.9 | 1.5 | 5.9 (3.4, 10.1) | (*20*) |
| Fold-reduction for Omicron relative to Delta | VFR | 10.0 | 10.0 | 3.9 (2.9, 5.5) | (*21*, *22*) |
| Titre relative to convalescent required to provide 50% protection from infection | $n_{i50}$ | 0.20 | 0.25 | 0.114 (0.064, 0.201) | (*13*) |
| Titre relative to convalescent required to provide 50% protection from severe disease | $n_{s50}$ | 0.03 | 0.25 | 0.029 (0.014, 0.058) | (*13*) |
| Titre relative to convalescent required to provide 50% protection from death | $n_{d50}$ | 0.03 | 0.25 | 0.018 (0.008, 0.038) | Assumed the same as hospitalisation |
| Shape parameter | k | 2.94 | 1 | 3.1 (2.5, 3.8) | (*13*) |
| Standard deviation of individual responses | - | 0.44 | Not fitted | - | (*13*) |
| Half-life of antibody decay: short (days) | $h_s$ | 58 | 5 | 48 (41, 55) | (*13*, *14*) |
| Half-life of antibody decay: long (days) | $h_l$ | 500 | 100 | 540 (370, 717) | (*15*) |
| Time period for switching (days) | $t_s$ | 90 | 20 | 94 (67, 121) | (*14*) |

The resulting dose-response curves relating NAT to vaccine efficacy are shown in **Figure S1**.

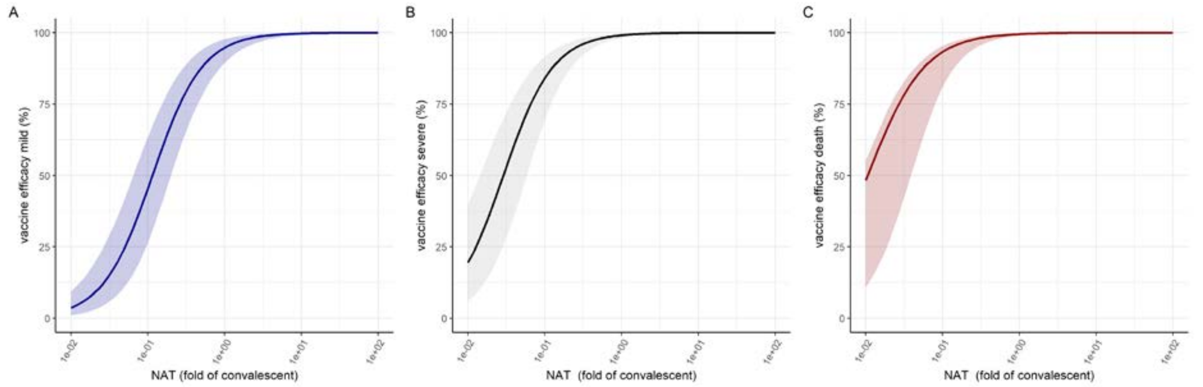


**Figure S1: Dose-response curves estimated from fitting to vaccine efficacy data** for the relationship between NAT (fold of convalescent) (x-axis) and vaccine efficacy against mild disease (A), severe disease (B) and death (C). The solid lines are the posterior median estimates and colour bands the 95% credible interval.

The model is fitted to one homologous regimen – PF primary and booster – and one heterologous regimen – AZ primary and PF booster. Data were estimates of vaccine efficacy at time points following either dose 1, dose 2 or the booster dose. We removed observations within the first 21 days post dose 1-, or 14-days post dose 2 or the boost and shifted the time respectively such that day 0 was either 21 or 14 days after the dosing, as our model does not capture the gradual increase in NAT over this short time-period.

We used the same process to capture infection-induced immunity derived from past infections. At each exposure, the NAT are assumed to be boosted by 1.7 with this parameter back calculated from the efficacy curves to give 90% protection against re-infection under our model as indicated from empirical studies. Each exposure or vaccine dose results in an additive increase in NAT. We imposed an upper limit of exp(5.0) for the total NAT an individual could achieve to represent an arbitrary biological limit.

For the forward simulations we assume that dose 2 is given 28 days following dose 1, whilst the booster dose is given 6 months following dose 2. As we only consider homologous vaccination in the simulations, we assumed that the boost for AZ is the same multiplier of AZ dose 2 as the estimated boost for PF is compared to PF dose 2.

#### *Population Model*

To explore the population impact of vaccines, we developed a stochastic, individual-based model (IBM) of SARS-CoV-2 transmission and vaccination ("safir"), including both vaccine-derived and infection-induced immune dynamics. The structure and epidemiology of the model broadly mirrors a previously published compartmental model (*37*) but implements these processes at the individual level to allow more flexible vaccine strategies to be implemented. The simulation updates on a discrete time step, but the specific size of the time step is under user control and can be made arbitrarily small, subject to computational constraints (we use a time-step of 0.2 days here). The model is implemented in R and C++, using the "individual" package to specify and run the simulation (*44*) and is open-source, with full details available at https://mrc-ide.github.io/safir/.

Within the IBM, each individual is assigned a 5-year age bin according to the demographics of the population, and transmission (force of infection) depends on a setting-specific age-structured contact matrix and also the baseline contact rate, which may be time-varying. In addition, infection from external (unmodelled) sources can be included to represent infectious contact with persons outside the modelled population (Figure S2). The states are:

- S = uninfected and therefore susceptible to infection
- E = exposed to infection but not yet infectious
- $I_{mild}$ = infected and infectious with mild symptomatic infection that does not require hospitalisation
- $I_{asymptomatic}$ = infected and infectious with asymptomatic infection that does not require hospitalisation
- $I_{case}$ = infected and infectious with disease that will require hospitalisation
- $I_{hosp}$ = individuals in hospital, which is comprised of:
  - o Mechanical ventilation = persons requiring mechanical ventilation
  - o Oxygen = persons requiring oxygen
  - o Neither = neither
- R = infections and cases that have recovered and are immune to re-infection
- D = cases that have died

Transitions between epidemiological states are summarised in **Figure S2**, with natural history parameters for SARS-CoV-2 infection, and age-stratified probabilities of requiring hospital care and the infection fatality ratio as reported in Hogan et al. (*37*).

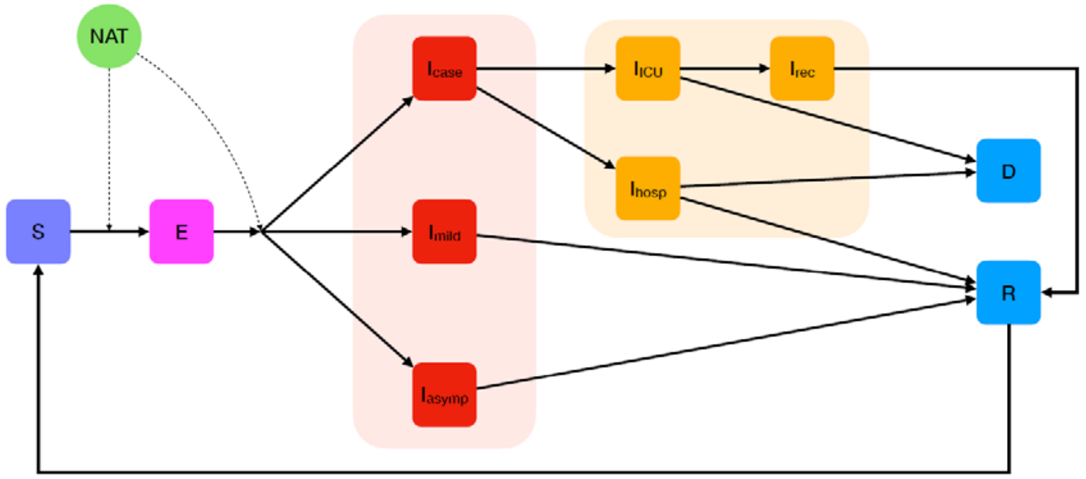


**Figure S2: Schematic representation of the compartmental epidemiology of the stochastic IBM**. The green circle denotes the neutralising antibody titres (NAT) for each individual which are tracked over time and influence the probability of being infected (from susceptible S to exposed E) and of developing disease requiring hospitalisation ($I_{case}$) given a breakthrough infection. $I_{mild}$ denotes mild symptomatic disease, $I_{asymp}$ asymptomatic infection, $I_{hosp}$ disease requiring hospitalisation, $I_{ICU}$ disease requiring ICU admission, $I_{rec}$ stepdown from ICU, D death, R recovered.

In addition, as described above, each individual stores a real-valued (floating point) log antibody titre value, which is altered by vaccination and infection. Upon receiving a vaccine dose, an individual's NAT is drawn from a vaccine-specific $log_{10}$-normal distribution and stored for that person. Each person's NAT decays linearly on a natural log scale, as described above.

In addition to vaccination, individuals receive an increase in NAT due to infection (instead of modelling temporary complete immunity as in previous compartmental models, where a person transitions into recovered "R" compartment where they dwell before returning to a susceptible "S" state). In the IBM, upon completion of the disease progression (when a person would have previously transitioned into the "R" compartment), the NAT derived from prior infection is added to that from vaccination (and vice versa) and stored for that individual, thereby capturing the interaction between past infection and vaccination (or vice versa). In order to capture different decay rates, the NAT from infection is stored separately.

Subsequent vaccine doses are correlated within individuals, meaning that subsequent antibody boosts from doses are equal to the random initial draw multiplied by the ratio of the previous mean dose-specific NAT to this dose-specific value. This allows us to implicitly capture individual level heterogeneity in immune response to vaccination. This new vaccine dose NAT is added to the current NAT for that individual. We further calculate the residual decayed NAT from the prior vaccine dose (calculated by subtracting the total decay on a natural log scale from the dose-specific logged NAT for the previous dose), and subtract this from the new vaccine dose NAT, so as to accurately implement the NAT profiles in Figure 1 within the IBM.

An individual's antibody titre is used to calculate protection as described above (capturing infection-induced immunity and vaccine-induced immunity in combination). We model protection against infection by reducing the probability that susceptible individuals transition to exposed, and protection against severe disease by reducing the probability of severe outcomes after being infected.

#### *Vaccine Allocation*

Vaccines are allocated according to an algorithm that accounts for available vaccine stock, the age-groups that are prioritised for this dose (and, optionally, future doses), minimum time delays between the receipt of subsequent doses, and coverage targets for each dose and for each age-group. The algorithm is similar to a previously published model (*37*) but generalised for an arbitrary number of doses and booster priority.

For each dose, we specify a matrix of coverage targets by age-group (columns) where rows are ordered prioritisation steps. Elements of the matrix thus identify for each step what target percentage dose coverage of that age-group needs to be fulfilled. We identify what dose is being distributed by distribution "phase" (e.g. on phase 2, the 2$^{nd}$ dose is being distributed). Within that distribution phase, the coverage target matrix specific to that dose/phase is used to distribute doses (subject to availability) according to each row of the matrix. When all coverage targets of a particular prioritisation step (a row) are fulfilled, the distribution algorithm moves to the next step (i.e. the next row). When coverage targets for all steps in a particular dose are achieved, we may move on to the next dose phase (i.e. distribute the next dose).

On each day for which there are at least some available doses, the algorithm checks which priority stage in a phase it is on. If a phase is completed, it advances to the next phase. If not, based on the targets for that step, the set of eligible persons to receive that dose is identified. This considers which age-groups have not yet met coverage targets, and which persons have had an adequate waiting period since their last dose (if phase > 1). Then doses are distributed to those individuals, subject to supply. If there are surplus doses remaining, the algorithm checks whether any age-groups are prioritised for phase + 1 booster doses and distributes those doses. If any doses remain, they are given out to age-groups in subsequent prioritisation steps. The algorithm stops if all targets for all doses are filled. The flowchart in **Figure S3** describes how the algorithm functions on a single day.

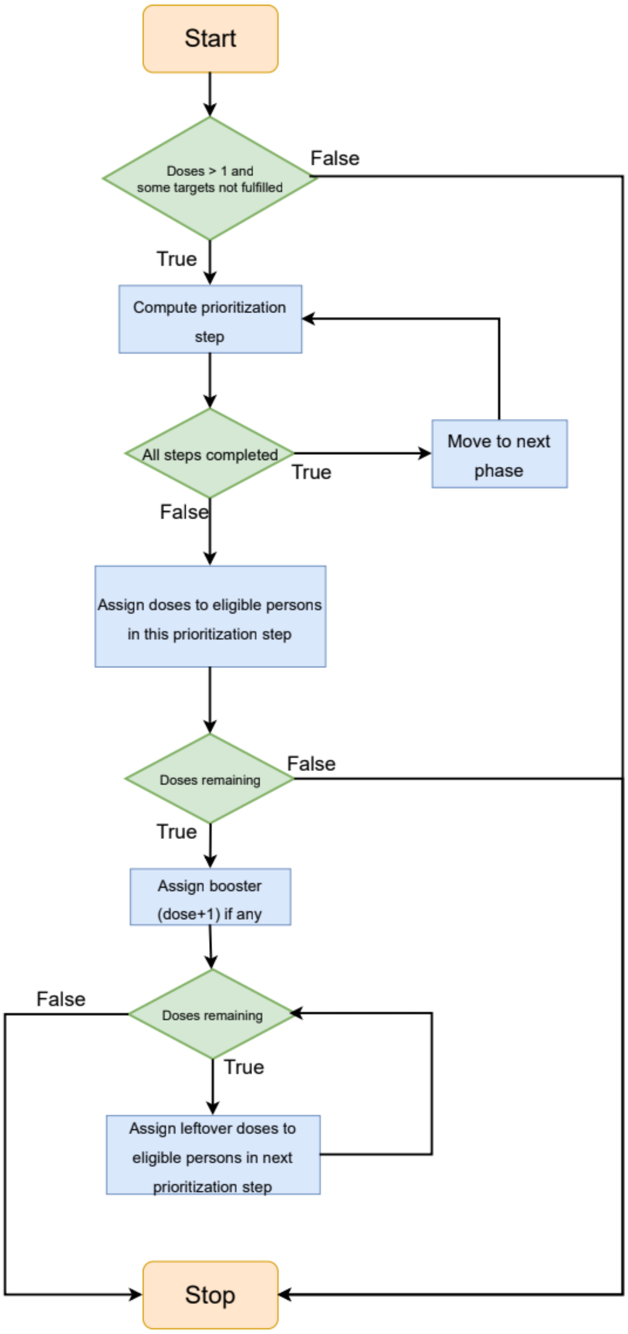


**Figure S3: Schematic diagram illustrating the COVID-19 vaccine allocation algorithm.**

### *Forward Simulations*

Following the methodology of previous work (*36*, *37*) we consider vaccine impact in four income settings: high-income, upper-middle-income, lower-middle-income, and low-income (HIC, UMIC, LMIC, and LIC, respectively), focussing on HIC and LMIC settings for the present analysis. We characterise each setting by contact pattern, using a typified contact matrix for each income setting (*36*, *37*) and demography, using the age distribution of the country with the median GDP in each income setting (HIC: Malta; LMIC: Nicaragua). We further consider that in LMIC countries, healthcare system capacity may be limited, and assume that once modelled hospitalised cases exceed a threshold in these settings, infected individuals experience worse outcomes, as in Walker et al (*36*). In HIC settings, we assume that healthcare systems are unconstrained.

Modelled transmission is allowed to vary over time by setting a time-varying reproduction number, $R_t$. Because the types and impacts of NPIs vary depending on setting, rather than modelling any specific NPIs (such as social distancing measures, mask-wearing, or school closures), we assume that the impact of these measures is reflected in the reproduction number. For the country settings in categories 1 and 2 (countries with substantial prior transmission before the rollout of vaccination), we construct a representative trajectory of transmission by varying $R_t$ for the Delta variant to match patterns observed in these settings (guided by country-level fits from our global model fitting – see [https://mrc-ide.github.io/global-lmic-reports](https://mrc-ide.github.io/global-lmic-reports)), such that an initial epidemic wave occurs between March and May 2020, a second wave occurs during December 2020 and January 2021. For the high-income countries we model a third wave (the Delta wave) occurring from September onwards (similar to that experienced in most European countries) with the resulting Rt for Delta reaching 3.5 whilst in the LMIC countries we allow an increase from mid-2021 to capture the earlier Delta waves with the resulting Rt for Delta reaching 4 Figure S4). This difference between HIC and LMIC likely represents the degree of ongoing NPIs in different countries. In an HIC setting, this initial trajectory results in approximately 840 deaths per million by the end of 2020, which aligns with the excess mortality experienced in high-burden high-income countries (*45*). For the country settings in category 3 (countries with minimal prior transmission before the rollout of vaccination), we construct a representative trajectory such $R_t < 1$ (no sustained transmission) until September 2021, at which point $R_t$ increases gradually to 3 for the Delta variant or from November 2021 to 3 (2.75, 3.25) for the Omicron variant (see below). We further explore scenarios in this category where transmission is suppressed until either November 2021, or April 2022, to allow different vaccination strategies to be implemented before NPIs are lifted (Figure 2A).

Our choice of Rt values for the Omicron variant was guided by comparison of the early growth rates of the Omicron epidemics in the UK and Australia. This is shown in **Figure S4**.

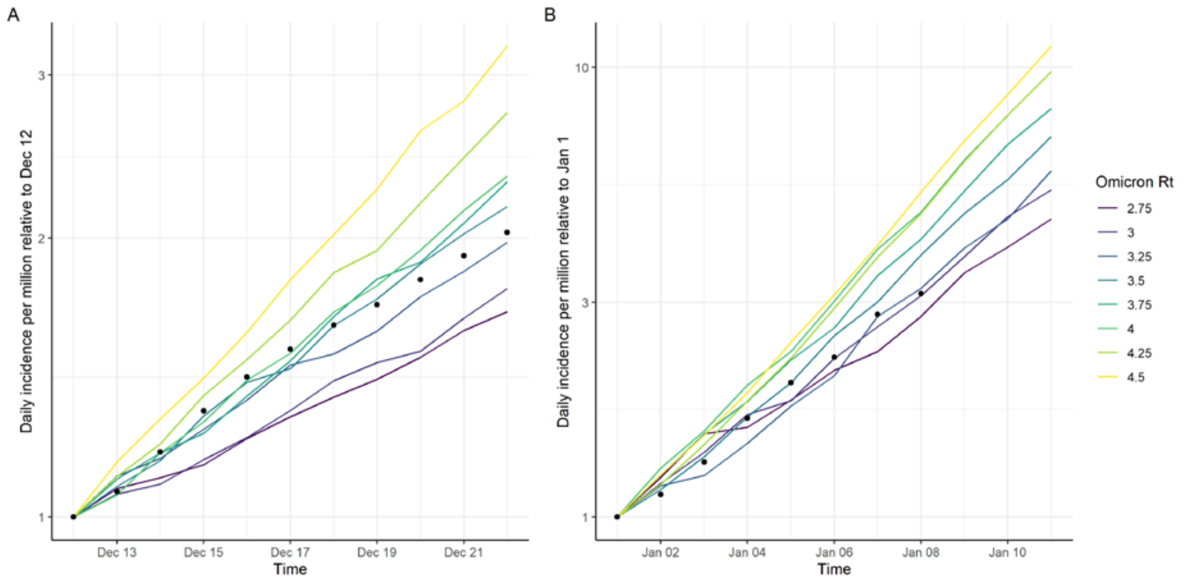


**Figure S4: Growth rates of the early phase of the Omicron wave in (A) the UK and (B) Australia.** Black points show the 7-day smoothed average of reported cases relative to the baseline date. Coloured lines show the simulated epidemic trajectories for different assumed levels of transmissibility of the Omicron variant.

For the analysis in category 1 (HIC countries), we model the rollout of vaccines to individuals starting at 80+ years, down to 10+ years, in consecutive 5-year age-groups, at a constant rate of 5% of the population receiving one dose per week, starting at 1 January 2021, with 90% uptake in each age-group. We then either cease to administer any additional doses ("10y+ 2 doses, no booster"), or administer booster doses of the same vaccine product to 60+ years individuals ("10y+ 2 doses, 60y+ booster"), at the same pace, 6 months following dose 2. We further simulate the rollout of booster doses down to 40+ years, 20+ years, and 10+ years.

For the analysis in category 2 (LMIC countries with partial vaccine coverage), we aim to consider the relative impact of administering doses to vaccinate the younger working-age population with their primary series, versus diverting those doses to vaccinate the older population with a booster dose. We therefore construct two scenarios. In the first, we commence vaccination from April 2021, delivering the first two doses to the 60+ years population, at a maximum constant rate of 2% of the population receiving one dose per week, and assuming 80% uptake and delivery of the AZ vaccine. Once this coverage is achieved, vaccination is paused until the delay between administration of the second and booster doses (6 months) is complete. For the first scenario, we then vaccinate the 60+ years population with booster doses, beginning with the oldest (80+ years) age-group. For the second scenario, we take the same number of doses as would be required to give boosters to 60+ years, deliver these doses to individuals younger than 60 years (2 doses per person), again working downwards through the population until that dose supply is exhausted. We construct these rollout scenarios such that the number of doses given out each day is equivalent between scenarios with the total number of vaccines available sufficient to allow 80% coverage with 3 doses for the 60+ population.

For the category 3 analysis (minimal prior transmission in HIC countries), we aim to quantify the impact of different strategies for lifting NPIs relative to vaccination coverage targets. We first simulated the impact of increasing $R_t$ (to $R_t = 4$) once 80% coverage is achieved with primary immunisation (2 doses, around August 2021). We consider the total infections and deaths if vaccination continues such that no boosters are administered; if boosters are then targeted to 60+ years; or if boosters are targeted to 10+ years. In all scenarios, the vaccination rate is constant at 5% of the population dosed per week. Second, we simulated the impact of increasing $R_t$ once the 60+ years population is boosted (around October 2021) and either stopping vaccination or continuing to deliver boosters down to 10+ year age-groups. Finally, we simulate the impact of increasing $R_t$ once boosters have been administered to the 10+ years population, assuming 90% uptake. We repeat these scenarios under the assumption that Omicron replaces Delta during November–December 2021; here we assume that if $R_t$ has already increased to $R_t = 4$, then $R_t$ further increases to 4.5. For the scenario where NPIs are not lifted until 2022, we assume a partial increase in $R_t$ to 3.5 during the period of Omicron replacement, and the remaining increase to $R_t = 4.5$ in March 2022.

To model the impact of Omicron upon transmission and immune escape, we consider scenarios where Omicron is more transmissible than the Delta variant consistent with the rapid replacement observed in South Africa and the UK. We introduce the Omicron variant gradually between 27/11/2021 and 31/03/2021. During this period we reduce fold reductions in antibody titres such that the estimated vaccine efficacy against circulating virus reaches the levels in Table 1 once Omicron has fully replaced Delta at 31/12/2021. We also increase the intrinsic transmission $R_t$ linearly over this period. To identify the appropriate level of $R_t$, we generated epidemics using $R_t$ values from 4 to 5.5 in steps of 0.25 from our HIC exemplar model and calculated the growth rate of the simulated epidemic between 12/12/21 and 21/12/21. We compared this to the growth rate of cases in the UK between the same dates (stopping as 21/12/21 to exclude reporting effects over the holiday period), finding that $R_t$=4.5 provided the closest match to the data. Finally, we incorporated a linear decrease in severity over the same time period, reducing the probability of requiring hospitalisation by 35% and the probability of severe disease (requiring ICU) by an additional 65%.

Assumptions about setting characteristics and vaccine rollout are summarised in **Table S2**. As sensitivity analyses, we further consider scenarios with a period of 12 months between dose 2 and the booster dose, and scenarios with a slower vaccine rollout.

**Table S2: Default scenarios and assumptions for the three broad modelled categories of country and epidemiological state.** Values in parentheses represent sensitivity analyses.

| | Income setting | HIC/UMIC | LMIC/LIC |
|---|---|---|---|
| Setting | Health system constraints | Unconstrained | Constraints present |
| | Contact patterns | HIC exemplar | LMIC exemplar |
| | Demography | HIC median | LMIC median |
| Vaccine characteristics | Product | Pfizer | Oxford-AstraZeneca |
| | Efficacy | See Table S1 | See Table S1 |
| | Short-term durability | Fitted (see Table S1) | Fitted (see Table S1) |
| | Long-term durability | Fitted (see Table S1) | Fitted (see Table S1) |
| Timing of roll-out and doses | Vaccine start date | 1 January 2021 | 1 April 2021 |
| | Timing of dose 2 | 28 days post-dose 1 | 28 days post-dose 1 |
| | Timing of booster dose | 180 days post-dose 2 (90 days) | 180 days post-dose 2 |
| | Vaccination rate | 5% per week (2.5% per week) | 2% per week (1.5% per week) |
| Vaccine targeting | Uptake | 90% | 80% |
| | Allocation | See Methods | See Methods |

Each model run was performed with a simulation size of one million, across 10 random seeds, and the median and upper and lower 95% credible intervals summarised from the raw model outputs. We used these outputs to calculate the daily number of infections and deaths from 1 February 2020 to 31 December 2022, for each of the scenarios considered, as well as the total numbers of infections and deaths from the beginning of vaccine rollout (1 January 2021) to the end of the simulation window.

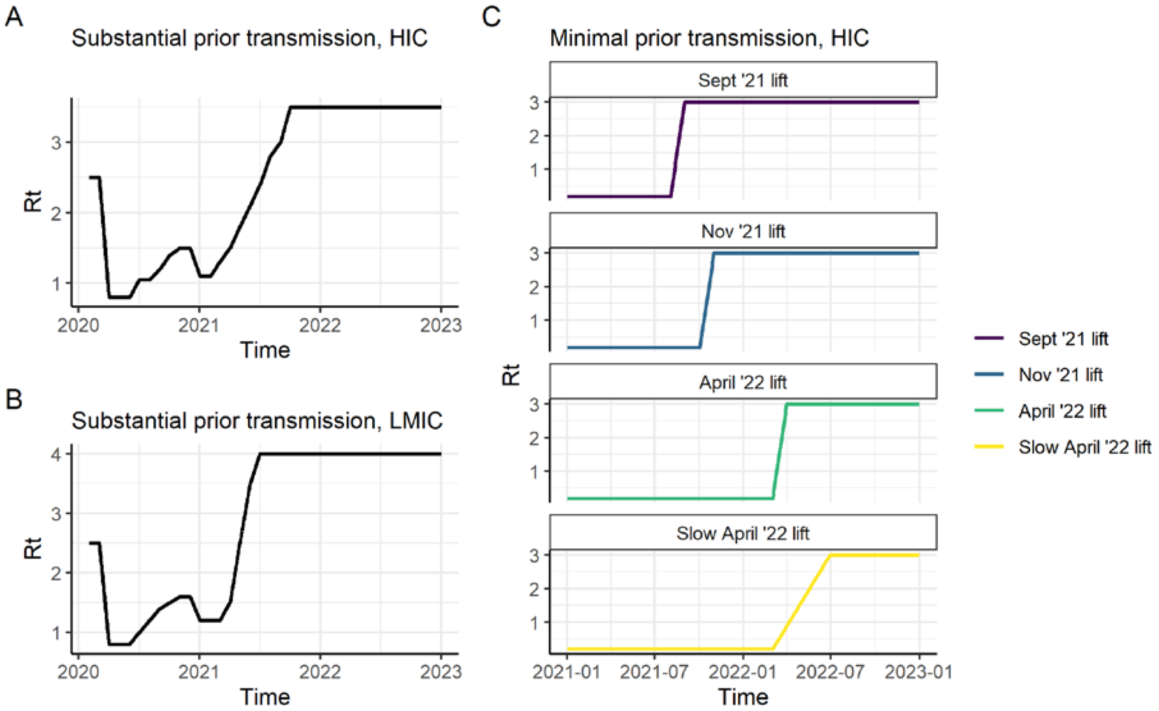


**Figure S5:** Modelled trajectories of the reproduction number $R_t$ over time for (A) the HIC setting with substantial prior transmission (category 1); (B) the LMIC setting with substantial prior transmission; and (C) the HIC setting with minimal prior transmission, for the four $R_t$ lifting scenarios (category 3).

### Supplementary Results

**Table S3: Sensitivity to estimated vaccine efficacy against mild disease, severe disease and death** for the AZ-PF and PF-PF vaccine regimens as a function of time since dose 2 or booster. Assumes that after a booster the decay rate is the same as that post dose 2. Estimates are shown for the Delta variant and the Omicron variant. Values shown are the posterior median and 95% credible intervals. pd2 = post dose 2; pb = post booster.

| Vaccine | Variant | 90d pd2 | 180d pd2 | 30d pb | 60d pb | 90d pb |
|---|---|---|---|---|---|---|
| *Efficacy against mild disease* | | | | | | |
| AZ-PF | Delta | 39.5 (38.6-40.4) | 23.7 (22.8-24.5) | 86 (85-87) | 80.5 (79-81.9) | 74.4 (72.5-76.3) |
| AZ-PF | Omicron | 9.5 (7.8-11.5) | 4.7 (3.8-5.8) | 49.5 (44.5-54.7) | 39.7 (35-44.9) | 31.8 (27.4-36.6) |
| PF-PF | Delta | 61.9 (60.9-62.8) | 43.6 (42.4-44.6) | 86 (85-87) | 80.5 (79-81.9) | 74.4 (72.5-76.3) |
| PF-PF | Omicron | 20.6 (17.2-24.4) | 11 (9-13.3) | 49.5 (44.5-54.7) | 39.7 (35-44.9) | 31.8 (27.4-36.6) |
| *Efficacy against severe disease* | | | | | | |
| AZ-PF | Delta | 80.5 (79.8-81.1) | 66.2 (65.2-67.2) | 97.5 (97.3-97.7) | 96.3 (95.9-96.6) | 94.8 (94.3-95.3) |

| | | | | | | |
|---|---|---|---|---|---|---|
| AZ-PF | Omicron | 39.7 (34.6-45.1) | 23.9 (20.1-28.1) | 86.1 (83.5-88.4) | 80.6 (77.2-83.7) | 74.6 (70.5-78.4) |
| PF-PF | Delta | 91.1 (90.7-91.5) | 83 (82.3-83.6) | 97.5 (97.3-97.7) | 96.3 (95.9-96.6) | 94.8 (94.3-95.3) |
| PF-PF | Omicron | 62.1 (56.7-67.2) | 43.9 (38.4-49.2) | 86.1 (83.5-88.4) | 80.6 (77.2-83.7) | 74.6 (70.5-78.4) |
| *Efficacy against death* | | | | | | |
| AZ-PF | Delta | 88.5 (88-89) | 78.6 (77.6-79.4) | 98.6 (98.5-98.8) | 98 (97.8-98.2) | 97.2 (96.9-97.5) |
| AZ-PF | Omicron | 55.2 (49.6-60.7) | 37 (31.8-42.3) | 92 (90.3-93.5) | 88.6 (86.3-90.6) | 84.6 (81.6-87.2) |
| PF-PF | Delta | 95 (94.8-95.3) | 90.1 (89.6-90.6) | 98.6 (98.5-98.8) | 98 (97.8-98.2) | 97.2 (96.9-97.5) |
| PF-PF | Omicron | 75.4 (70.8-79.3) | 59.3 (53.6-64.5) | 92 (90.3-93.5) | 88.6 (86.3-90.6) | 84.6 (81.6-87.2) |

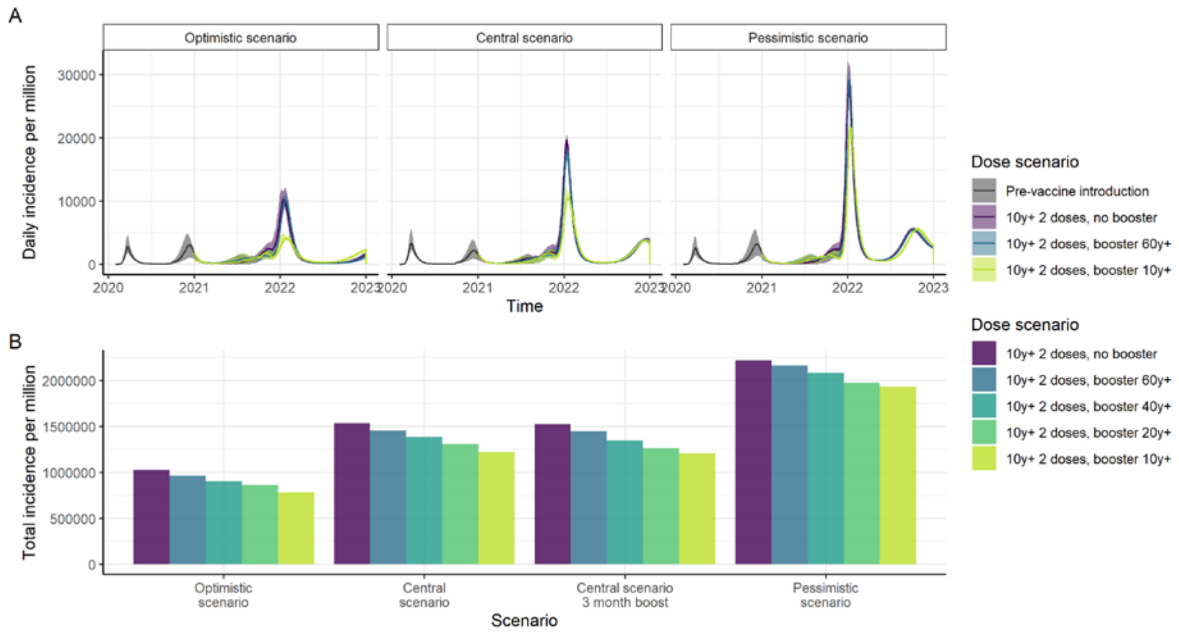


**Figure S6: Impact of vaccination (incidence) in a high-income country setting with substantial prior transmission and high vaccine access**. A) Daily incidence per million for the central, optimistic and pessimistic scenarios. B) Total incidence per million from vaccine introduction at the beginning of 2021 through to the end-2022 for the three scenarios assuming Omicron replaces Delta during November–December 2021, and assuming a 6-month period between dose 2 and the booster, with an additional scenario showing the impact of assuming a 3-month period. Vaccine impact is shown for five scenarios: no booster doses, booster doses to those aged 60+, booster doses to those aged 40+, booster doses to those aged 20+ and booster doses to those aged 10+. Impact is shown for three epidemiological scenarios: central, with our central estimate of the VFR (3.9), Omicron Rt=3.5 (the same as Delta) and the risk of hospitalisation reduced by 50% for Omicron compared to Delta; optimistic with our lower estimate of VFR (2.9), Omicron Rt=3.25 and risk of hospitalisation reduced by 70%; and pessimistic with our upper estimate of the VFR (5.5), Rt=3.75 and risk of hospitalisation reduced by 40%.  All scenarios assume 90% vaccine uptake in each group and the risk of death further reduced by 58% from the risk of hospitalisation.

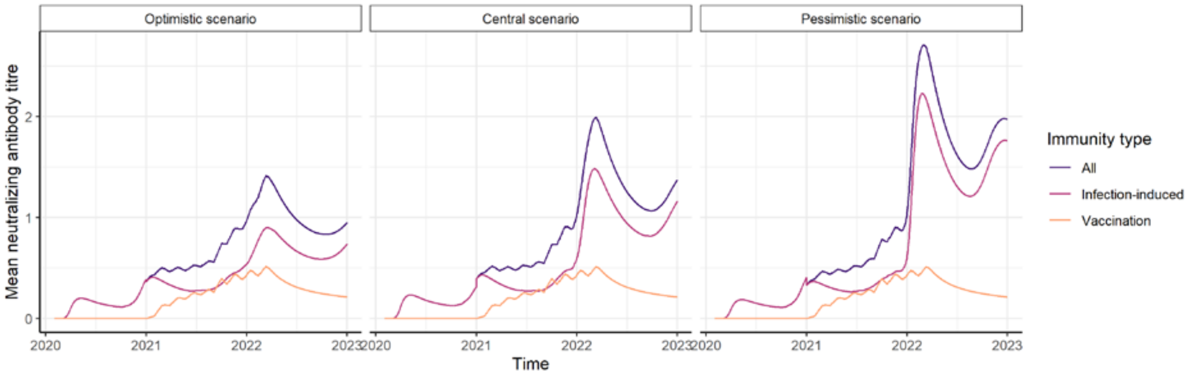


**Figure S7: Mean infection-induced (pink), vaccine-induced (orange), and total (purple) neutralizing antibody titres (NAT) over time.** Results are shown for a high-income country setting with substantial prior transmission and high vaccine access. Estimates are shown for three epidemiological scenarios: central, with our central estimate of the VFR (3.9), Omicron Rt=3.5 (the same as Delta) and the risk of hospitalisation reduced by 50% for Omicron compared to Delta; optimistic with our lower estimate of VFR (2.9), Omicron Rt=3.25 and risk of hospitalisation reduced by 70%; and pessimistic with our upper estimate of the VFR (5.5), Rt=3.75 and risk of hospitalisation reduced by 40%. All scenarios assume 90% vaccine uptake in each group and the risk of death further reduced by 58% from the risk of hospitalisation.

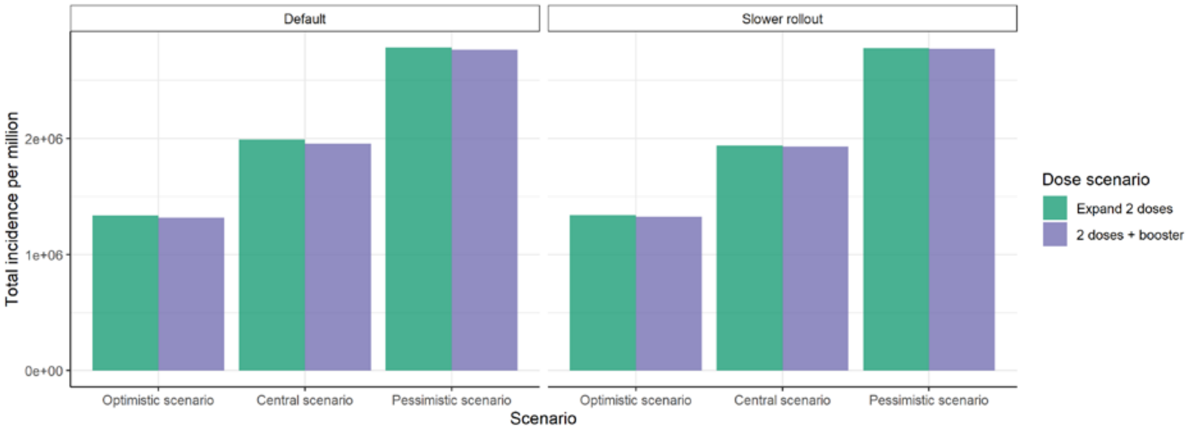


**Figure S8: Impact of vaccination on total incidence in a low-middle-income country setting with substantial prior transmission and low vaccine access**, assuming the population 40+ years is initially targeted. Two strategies for distributing a fixed limited supply are shown. In the first ("Expand 2 doses"), no booster doses are administered, and the supply is therefore delivered to a wider proportion of the population. In the second ("2 doses + booster") the same supply is delivered to the 40+ age-group (2 dose primary immunisation and booster dose 6 months post dose 2) and no younger groups receive the primary series. The bars show total incidence per million population from vaccine introduction in 2021 to end-2022.Total incidence is shown for the default vaccine rollout (2% of the population per dose per week) or a slower rollout (1% per week, "Slower rollout"). Impact is shown for three epidemiological scenarios: central, with our central estimate of the VFR (3.9), Omicron Rt=4 (the same as Delta in this setting) and the risk of hospitalisation reduced by 50% for Omicron compared to Delta; optimistic with our lower estimate of VFR (2.9), Omicron Rt=3.75 and risk of hospitalisation reduced by 70%; and pessimistic with our upper estimate of the VFR (5.5), Rt=4.25 and risk of hospitalisation reduced by 40%.  All scenarios assume 80% vaccine uptake in each group and the risk of death further reduced by 58% from the risk of hospitalisation.

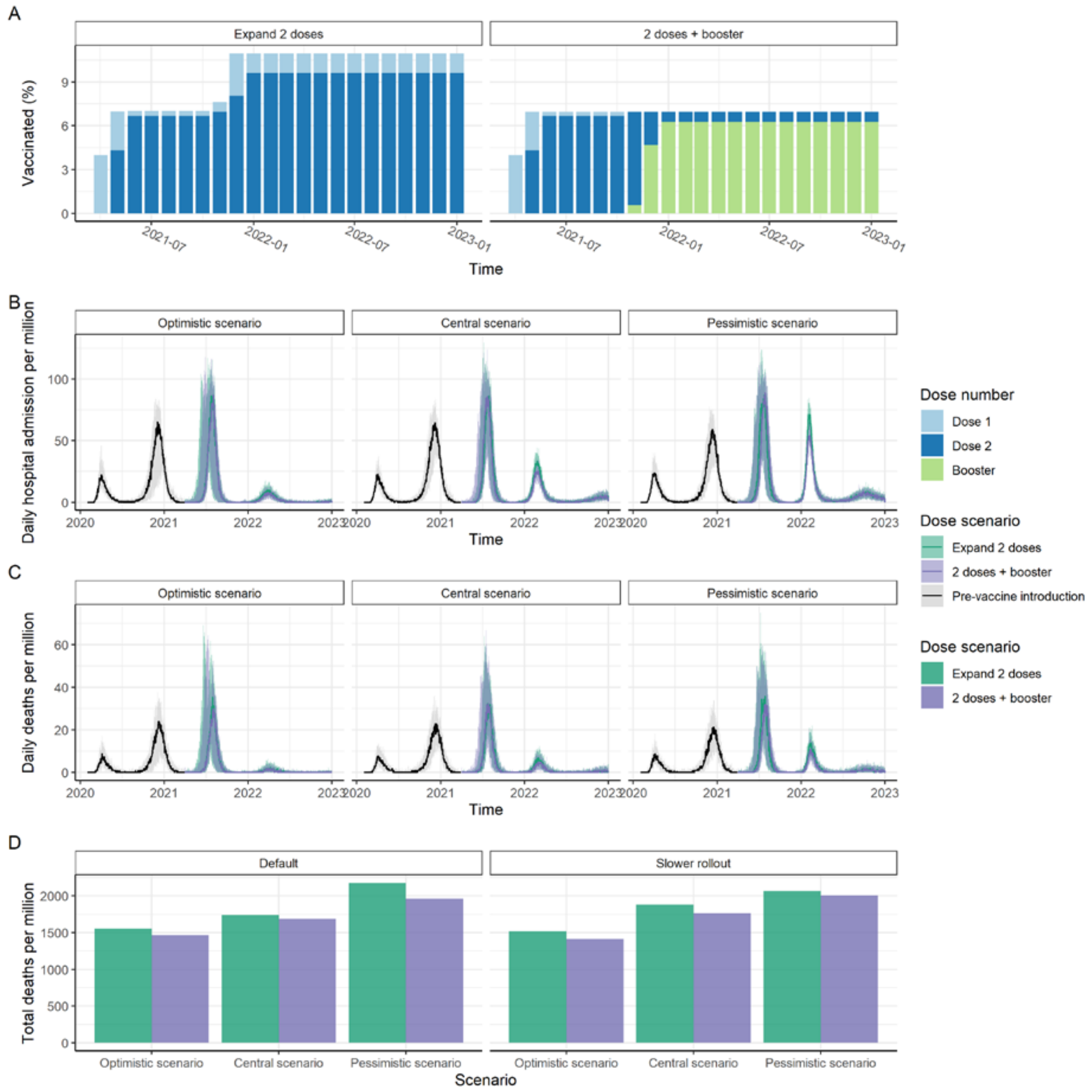


**Figure S9: Impact of vaccination in a low-middle-income country setting with substantial prior transmission and low vaccine access, where individuals 60+ years are initially targeted.** Two strategies for distributing a limited vaccine supply are shown (assuming AZ is used for both the primary and booster doses). In the first (“Expand 2 doses”), no booster doses are administered, and the supply is therefore delivered to a wider proportion of the population. In the second (“2 doses + booster”) the same supply is delivered to the 40+ age-group (2 dose primary immunisation and booster dose 6 months post dose 2) and no younger groups receive the primary series. (A) Cumulative proportion of the population receiving dose 1 (light blue), dose 2 (dark blue) and the booster (green) each month. (B) Daily hospital admissions; and C) Daily deaths per million population for our central, best-case and worst-case scenarios (see legend to Figure 2). Total deaths per million population from the start of vaccination in 2021 to end-2022 for (D) the three scenarios with the default and slower roll-out rate. Impact is shown for three epidemiological scenarios: central, with our central estimate of the VFR (3.9), Omicron Rt=4 (the same as Delta in this setting) and the risk of hospitalisation reduced by 50% for Omicron compared to Delta; optimistic with our lower estimate of VFR (2.9), Omicron Rt=3.75 and risk of hospitalisation reduced by 70%; and pessimistic with our upper estimate of the VFR (5.5), Rt=4.25 and risk of hospitalisation reduced by 40%. All scenarios assume 80% vaccine uptake in each group and the risk of death further reduced by 58% from the risk of hospitalisation. Total deaths are shown for the default vaccine rollout (2% of the population per dose per week) and booster dose scenario (180 days post dose 2, “Default”), as well as assuming a slower rollout (1% per week, “Slower rollout”).

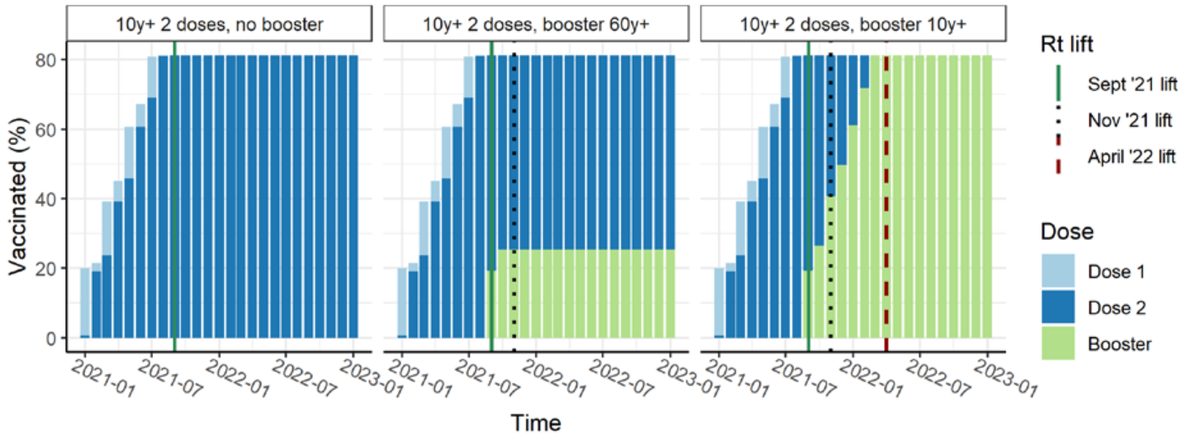


**Figure S10: Percentage of the population vaccinated with either one dose (light blue bars), two doses (dark blue), or two doses plus a booster dose (light green), per month from January 2021 for the high-income country setting with high vaccine access and minimal prior transmission.** The vertical lines show the different options for lifting NPIs (i.e. increasing $R_t$) relative to the rollout of vaccine doses. The “Sept ’21 lift” corresponds to lifting NPIs after 80% of the population has received two doses; “Nov ’21 lift” corresponds to lifting following rollout of booster doses to the 60+ years population; and “April ’22 lift” corresponds to lifting after boosters have been rolled out to the 10+ years population. The population health impact for these roll-out and lifting scenarios is in Figure 4.

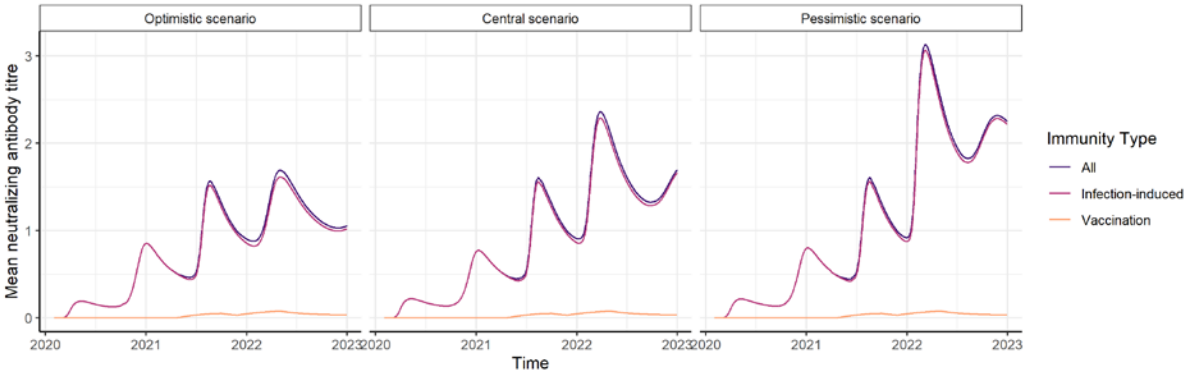


**Figure S11: Mean infection-induced (pink), vaccine-induced (orange), and total (purple) NAT over time in a high-income country setting with minimal prior SARS-CoV-2 transmission and high vaccine access**. Results are shown for the central scenario. All scenarios assume 90% vaccine uptake in each group.

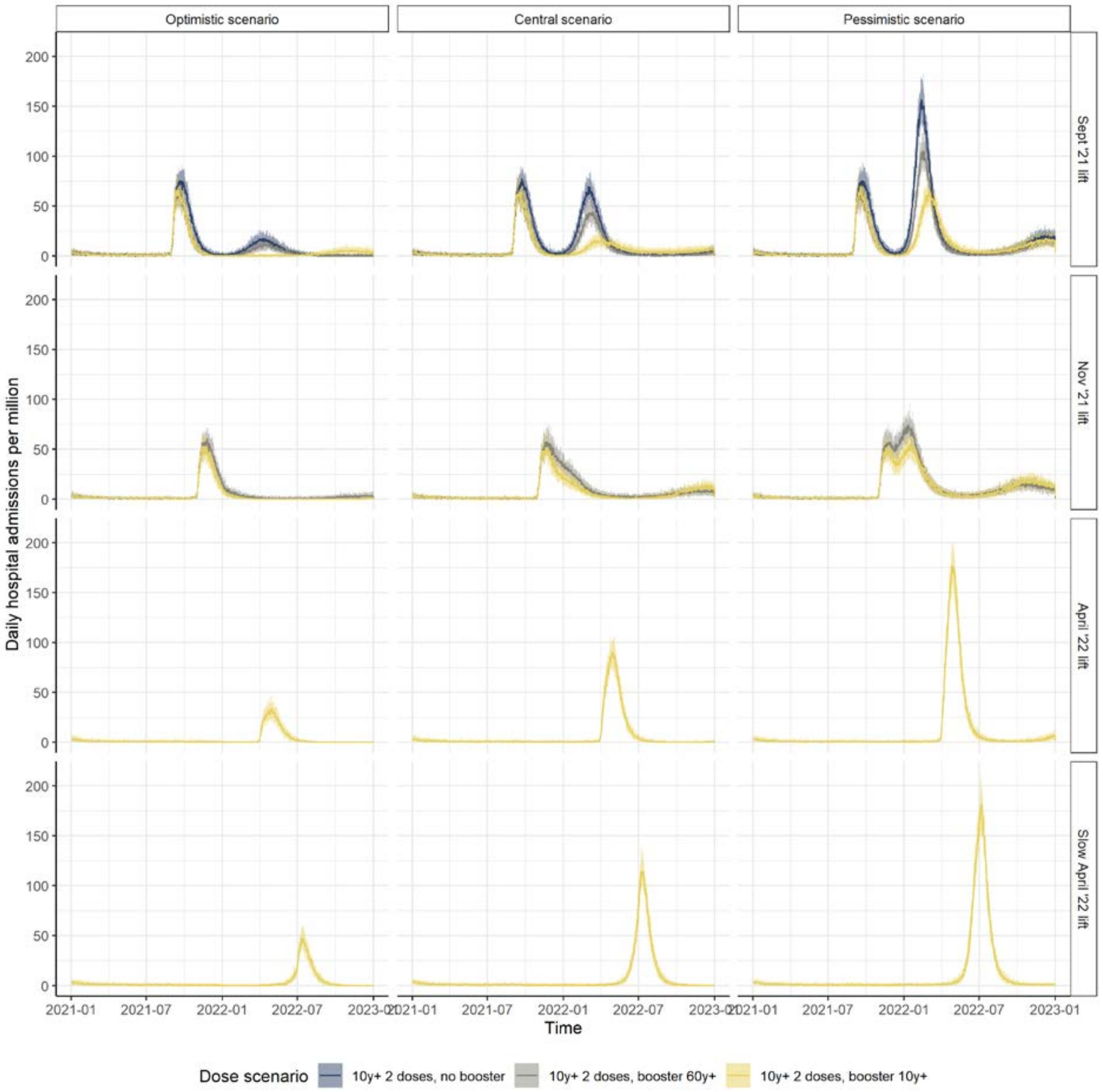


**Figure S12: Impact of vaccination in a high-income country setting with minimal prior SARS-CoV-2 transmission and high vaccine access.** Daily hospital admissions per million population for the scenario where 80% of the population is immunised (2 doses) before NPIs are lifted (i.e. $R_t$ increases). Following Rt increasing ("Sept '21 lift"), vaccination continues at a population dose rate of 5% per week until 90% uptake is achieved in the 10+ years population (brown); boosters are administered to the 60+ years population 6 months post dose 2 (dark blue); or boosters are administered to the 10+ years population (yellow). Additional panels show the impact of lifting NPIs after boosters are given to 60+ years ("Nov '21 lift") or after boosters are given to 10+ years ("April '22 lift" representing a lifting over a one-month period; "Slower April '22 lift" representing lifting over a four-month period). Impact is shown for three epidemiological scenarios: central, with our central estimate of the VFR (3.9), Omicron Rt=3 (the same as Delta) and the risk of hospitalisation reduced by 50% for Omicron compared to Delta; optimistic with our lower estimate of VFR (2.9), Omicron Rt=2.75 and risk of hospitalisation reduced by 70%; and pessimistic with our upper estimate of the VFR (5.5), Rt=3.25 and risk of hospitalisation reduced by 40%. All scenarios assume 90% vaccine uptake in each group and the risk of death further reduced by 58% from the risk of hospitalisation.
